# A digital health intervention to support patients with chronic pain during prescription opioid tapering: a pilot randomised controlled trial

**DOI:** 10.1101/2023.05.10.23289771

**Authors:** Ali Gholamrezaei, Michael R Magee, Amy G McNeilage, Leah Dwyer, Alison Sim, Manuela L Ferreira, Beth D Darnall, Timothy Brake, Arun Aggarwal, Meredith Craigie, Irina Hollington, Paul Glare, Claire E Ashton-James

## Abstract

**Introduction:** Recent changes in opioid prescribing guidelines have led to an increasing number of patients with chronic pain being recommended to taper. However, opioid tapering can be challenging, and many patients require support.

**Objectives:** We evaluated the feasibility, acceptability, and potential efficacy of a co-designed psycho-educational video and SMS text messaging intervention to support patients with chronic pain during prescription opioid tapering.

**Methods:** A pilot randomised controlled trial was conducted. In addition to their usual care, participants in the intervention group received a psycho-educational video and 28 days of text messages (two SMS/day). The control group received usual care. The feasibility, acceptability, and potential efficacy of the intervention were evaluated. The primary outcome was opioid tapering self-efficacy. Secondary outcomes were pain intensity and interference, anxiety and depression symptom severity, pain catastrophising, and pain self-efficacy.

**Results:** Of 28 randomised participants, 26 completed the study (13 in each group). Text message delivery was 99.2% successful. Most participants rated the messages as useful, supportive, encouraging, and engaging, 78.5% would recommend the intervention to others, and 64% desired a longer intervention period. Tapering self-efficacy (Cohen’s *d* = 0.74) and pain self-efficacy (*d* = 0.41) were higher and pain intensity (*d* = 0.65) and affective interference (*d* = 0.45) lower in the intervention group at week 4.

**Conclusions:** It is feasible, acceptable, and potentially efficacious to support patients with chronic pain during prescription opioid tapering with a psycho-educational video and SMS text messaging intervention. A definitive trial has been initiated to test a 12-week intervention.

## INTRODUCTION

Chronic pain is a leading cause of disability worldwide.^8^ Opioid medications are commonly prescribed for managing chronic non-cancer pain (CNCP).^7^ However, accumulating evidence has revealed limited benefits and dose-related harms associated with long-term opioid therapy (LTOT), promoting changes to prescribing guidelines and regulations.^14^ As a result, patients with CNCP are increasingly being advised to gradually taper off LTOT or reduce their opioid dose under clinical supervision.^9^

Tapering LTOT poses challenges for both patients and clinicians.^24, 36^ Patients often express concern about increased pain when considering tapering.^39^ Dose reduction can lead to unpleasant withdrawal symptoms and negatively impact mood and pain.^24, 36^ Recent studies have also raised concerns about the potential risk of overdose and harms associated with opioid tapering,^2^ underscoring the need for additional support during this process.^4^ Indeed, access to a range of supports, including pain education, monitoring, a strong patient-physician relationship, and strategies for managing pain and withdrawal symptoms, has been found to shape the trajectory of patients’ tapering experience.^13, 19, 36, 38^ However, access to support for opioid tapering remains a pervasive challenge.^18, 24, 26^

Digital health technologies using mobile phones (mHealth) are emerging as a solution to the global challenge of providing patients with access to support for health behaviour change.^16, 34^ These technologies can be cost-effective in delivering healthcare services on a large scale, especially when addressing chronic conditions, and can be adapted to the needs of diverse demographic groups and health conditions.^16, 22, 40^ Available evidence suggests that digital health interventions can improve pain interference and severity, psychological distress, and health-related quality of life in people with chronic pain.^43^ Evidence for the effectiveness of digital health interventions to support patients with CNCP during tapering LTOT is promising but limited.^4^

Our previous research has shown that patients with CNCP generally have positive attitudes toward using digital health technologies, particularly Short Message Service (SMS) text messages, to support them with opioid tapering.^33^ Studies have also found that educational videos can be an effective means of providing patients with information about chronic pain, pain self-management, and opioid tapering.^12, 15, 23^ Educational videos have been shown to increase the opioid-tapering self-efficacy of people who are currently on LTOT for chronic pain.^15^ Building on this foundational research, we co-designed a mobile health intervention, consisting of a brief psycho-educational video and SMS text messaging, for patients with CNCP who are tapering prescription opioids under clinical supervision.^32^ Both patients and clinicians have rated this program as appropriate, useful, and likely to be effective in supporting patients during opioid tapering.^32^

The primary objectives of this pilot trial were (1) to assess the acceptability of an mHealth intervention designed to improve opioid tapering self-efficacy in patients with CNCP, and (2) to evaluate the feasibility of the intervention and the methodology for a future definitive trial. The secondary objectives of this trial were (1) to evaluate the potential efficacy of the intervention and (2) to obtain estimates that can be used to design a future definitive trial.

## METHODS

Full details of the study methods are described in the published study protocol.^31^ The study was approved by the Northern Sydney Local Health District Research and Ethics Committee (ID number 2020/ETH03288) and pre-registered with the Australian New Zealand Clinical Trials Registry (registration number ACTRN12621000795897).

### Trial design and study setting

The study was a pilot, single-blind randomised controlled trial (RCT) with two parallel arms (intervention and control group), allocated in a 1:1 ratio. Recruitment was open at outpatient multidisciplinary pain management clinics located in three public hospitals in Australia (in Sydney and Adelaide).

### Participants and recruitment

Participants in this study were individuals with CNCP who were tapering their opioids under the supervision of a clinician. Eligible participants were identified by the clinicians based on the criteria listed in Table 1. Those who expressed interest in the study were contacted by a research team member (MM) to confirm eligibility^1, 11, 17^ and provided detailed study information including explaining randomisation process and what each study group will receive during the study period.

**Table 1.** Eligibility criteria.

|  |
| --- |
| Inclusion criteria<br>- Age 18 years or older.<br>- Diagnosed with a chronic (>3 months) pain condition.<br>- Have been using opioid analgesics at a dose of at least 40 mg/day oral morphine equivalent for at least 4 weeks.<br>- Have been advised by a clinician to taper opioids.<br>- Were tapering opioid medications voluntarily, as indicated by verbalised willingness and consent.<br>- Have been tapering or would be tapering their opioid medications at the time of enrolment.<br>- Able to understand written and spoken English.<br>- Own a mobile phone that receives SMS text messaging.<br>- Able to give written informed consent and comply with study procedures.<br>Exclusion criteria<br>- Cognitive impairment or intellectual disability.<br>- Evidence of severe opioid use disorder.<sup>1</sup> Illicit substance use was not an exclusion criterion unless indicating a severe opioid use disorder.<br>- History of primary psychotic disorder, bipolar affective disorder, bipolar disorder with psychotic features, depressive disorder with psychotic features, borderline personality disorder, antisocial personality disorder or positive family history (first-degree relative) of psychotic disorder or bipolar affective disorder such that participants might be at more than low/negligible risk by participating in the study.<br>- Any other major, poorly controlled medical or mental health comorbidity.<br>- Participation in another clinical trial concurrently. |

### Treatment Groups

#### Usual care

In this study, ‘usual care’ was defined as the care provided at the pain clinics by a multidisciplinary team of specialist pain medicine physicians, clinical psychologists, physiotherapists, and nurses. The decision to taper and the tapering schedule were negotiated between the patient and their physician. Participants randomised to the usual care group did not receive the mHealth support intervention.

#### Usual care + mHealth support intervention

The development of the intervention is described in detail elsewhere.^32^ The psycho-educational video and library of text messages were co-designed with consumers and clinicians.^32^ The 10-minute video provided information about pain, opioid tapering, and pain self-management strategies as well as socio-emotional support in the form of testimonials. The content of the text messages reinforced the content of the video.

The mHealth intervention was provided in addition to the usual care given at the pain clinics. Participants randomised to the intervention group were given a video embedded in the Research Electronic Data Capture (REDCap) software,^21^ and the link was sent to their email. They then received two text messages per day (mid-morning and mid-afternoon) for 28 days. The messages were standardised in content and delivery across participants but were semi-personalised using the recipient’s first name in some of the messages. Messages were sent using a commercial software (Message Media, Message4U Pty Ltd, Melbourne, Vic). Participants were informed that the text messages were one-way. However, they could reply with “STOP” to a text message if they did not want to receive further messages.

### Allocation and Blinding

After completing the baseline assessments, participants were randomised to the study groups by the REDCap software, ensuring allocation concealment. Participants were informed of their group allocation via email (see the study protocol for the details^31^). Treating clinicians were blinded to the study arms. Participants completed questionnaires online. If necessary, data collection by phone call was performed by data collectors who were blinded to the study arms.^35^ The statistician was also blinded to the study arms.

### Outcome measures

The full list of the study (outcome) measures and assessment timeline are provided in the Supplementary Materials (Table S1). Self-reported survey data were collected online using REDCap software.

#### Acceptability and feasibility measures

Participants in the intervention group were surveyed to obtain their feedback on the acceptability and feasibility of the intervention. The feedback survey (Supplementary Materials) included rating scales and open-text responses to capture the likelihood of recommending the intervention to others, perceived usefulness of the intervention, levels of engagement with the intervention, barriers of engagement, messages readability, and preferred frequency and timing of the messages.^20, 45^ Feasibility was assessed by evaluating the delivery of the messages sent, numbers and reasons for exclusions and dropouts, as well as questionnaires completion and missing data rates.

#### Potential efficacy measures

The potential efficacy of the intervention was measured using a one-item scale of general self-efficacy to taper prescription opioids (OTSEQ, Opioid-Tapering Self-Efficacy Questionnaire). The OTSEQ was developed using Bandura’s self-efficacy theory and guides for constructing self-efficacy scales (Supplementary Materials).^6^ The scale asked participants to rate their confidence in reducing their dose of opioid medication by selecting a number from 0 (‘not at all confident’) to 100 (‘completely confident’). Face validity was evaluated by interviewing clinicians and patients with CNCP who had experienced opioid tapering. In addition, we measured pain intensity and interference using the three-item Pain, Enjoyment of Life and General Activity scale (PEG)^28^ and measured mood using the Generalised Anxiety Disorder 2-item^30^ and the Patient Health Questionnaire-2.^29^ These outcomes were measured at baseline and then every week for four weeks. Tapering self-efficacy was also measured in the intervention group immediately after watching the video.

Opioid dose and its change were assessed by asking participants to report their medications at baseline, and each week during the trial participants explained any changes in their opioid medication over the past week using an open-ended question. Total daily opioid use was converted to mg of oral morphine equivalents.^5^ Participants were asked weekly if they had experienced any withdrawal symptoms or felt unwell in the past week. Accordingly, the cumulative incidence of withdrawal symptoms over the trial period was measured. Pain catastrophising was measured using the six-item Concerns about Pain Scale (CAP-6).^3^ Pain self-efficacy was measured using the 10-item Pain Self-Efficacy Questionnaire (PSEQ).^37^ Pain catastrophising and self-efficacy were measured at baseline and then at week 4. Participants also rated their level of satisfaction with the care they received over the past 4 weeks using a 7-point Likert scale ranging from ‘very dissatisfied’ to ‘very satisfied’.^11^

#### Exploring factors associated with opioid tapering self-efficacy

We explored some factors that could potentially be associated with opioid tapering self-efficacy including readiness to taper,^11^ patient–physician relationship,^44^ perceived social support^27^, expectations regarding changes in pain and mood after opioid tapering, and autonomy (perceived degree of choice, see Supplementary Materials for the related scales).

### Statistical analysis

#### Sample size

To assess whether the intervention was acceptable to 70% of the participants with a 20% precision rate, 18 participants were needed in the intervention arm. To evaluate the potential efficacy of the intervention, 12 participants were needed in each group assuming a medium standardised effect size (Cohen’s *d* = 0.5) and using the 80% one-sided confidence interval (CI) approach, which is recommended for pilot trials.^10^ Therefore, the sample size was set at 20 participants for each study arm to address the study objectives and assuming a 10% loss to follow-up during the study period (see Supplementary Materials for more details).

#### Statistical methods

Descriptive statistics were used for reporting feasibility and acceptability measures. The linear mixed-effects model was used to analyse outcomes of potential efficacy. The main effect of the group was tested to estimate the overall difference in outcomes between the two groups across all timepoints (weeks 1 to 4). Then, group-by-time interaction was included in the model to assess if and how the effect might have changed over time. Pairwise contrasts were done to explore comparing the outcomes between the two groups at each week of the study. According to the pre-registered analysis plan,^31^ we used the one-sided 80% CI method in this pilot study. With this approach, we were interested in whether the difference estimates were larger or smaller than zero (depending on the predicted direction of the effect), and did not aim to formally undertake hypothesis testing procedures to prove the efficacy of the intervention.^10^ Hence, no correction was done for multiple comparisons for this pilot study. Data are presented as difference estimates and one-sided CI80%. Cohen’s *d* (effect size) was calculated based on the estimates and standard errors. All analyses were done using SAS software (V.9.4, see Supplementary Materials for more details).

## RESULTS

### Recruitment

Recruitment was open from August 2021 to November 2022. Thirty-nine potential participants were referred from the three study sites. Of these, 28 (72%) were eligible, consented, and enrolled in the study. Recruitment was stopped before reaching the planned sample size of 40 as it exceeded the available funding period. All enrolled participants were randomised. Following randomisation, one participant from each group dropped out of the study (loss to follow-up). In total, 13 participants in each group completed the study (Figure 1). All participants were included in an intention-to-treat analysis. The mean age (±standard deviation) was 50 (±12) years and 19 were female. Duration of pain and opioid therapy (median [25%, 75% quartiles]) were 10 (6, 23) and 6 (3, 10) years, respectively. There were no significant differences between the two groups in demographic or baseline characteristics (Table 2).

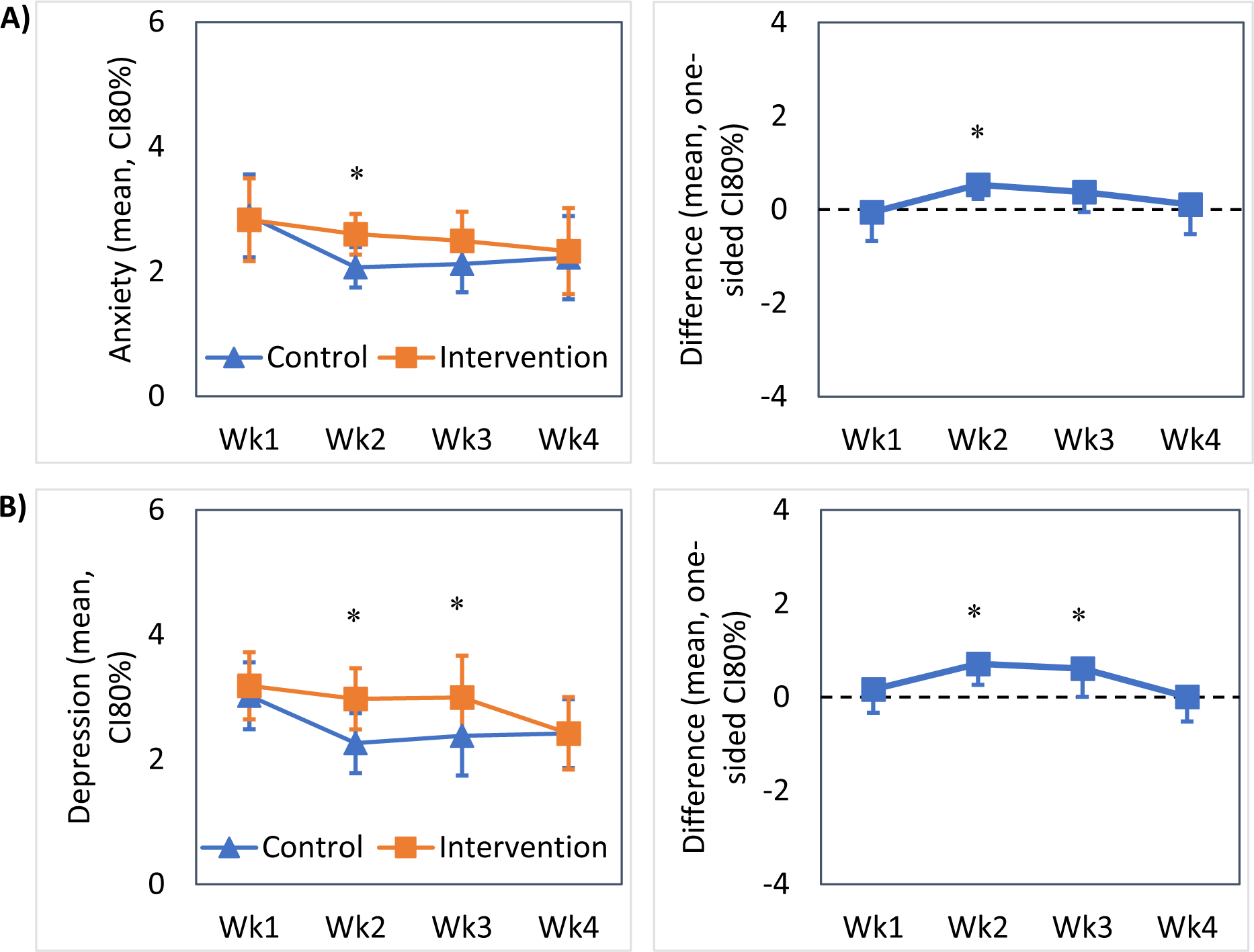

**Table 2.** Comparison of baseline and demographic characteristics between the two groups.

|  |  |  |  |
| --- | --- | --- | --- |
|  | Control<br>n = 14 | Intervention<br>n = 14 | P value |
| Age, year | 49.5 ± 13.8 | 50.5 ± 10.3 | 0.830* |
| Female | 10 (71.4) | 9 (64.2) | >0.999† |
| Education¥ |  |  | 0.212‡ |
| High school | 4 (30.7) | 3 (21.4) |  |
| Vocational | 7 (53.8) | 5 (35.7) |  |
| University | 2 (15.3) | 6 (42.8) |  |
| Employed¥ | 6 (42.8) | 4 (28.5) | 0.694† |
| Married/Domestic partnership¥ | 6 (42.8) | 7 (50) | >0.999† |
| *Place of usual residence* |  |  | >0.999† |
| Metropolitan | 10 (71.4) | 9 (64.2) |  |
| Regional | 2 (14.2) | 3 (21.4) |  |
| Rural | 2 (14.2) | 2 (14.2) |  |
| Number of other people in the household | 1.6 ± 1.2 | 1.3 ± 1.2 | 0.539* |
| *Pain conditions* |  |  |  |
| Neuropathic pain | 8 (57.1) | 6 (42.8) | 0.706† |
| Arthritis | 8 (57.1) | 4 (28.5) | 0.251† |
| Back/Neck pain | 7 (50) | 10 (71.4) | 0.440† |
| Other pain conditions | 8 (51.7) | 10 (71.4) | 0.694† |
| Number of pain conditions | 2.5 [1.2, 3.7] | 3 [2, 3.7] | 0.925‡ |
| Pain duration, years | 10 [6, 22] | 12.5 [6, 28] | 0.942‡ |
| *Comorbidities* |  |  |  |
| Psychiatric/Mental Health conditions | 12 (85.7) | 11 (78.5) | >0.999† |
| Cardiovascular | 5 (35.7) | 6 (42.8) | >0.999† |
| Respiratory | 3 (21.4) | 5 (35.7) | 0.677† |
| Endocrinologic | 3 (21.4) | 5 (35.7) | 0.677† |
| OME, mg/day | 118 [99, 194] | 105 [69, 195] | 0.593‡ |
| Duration of LTOT, year | 6 [2, 10] | 6.5 [4.1, 9.9] | 0.846‡ |
| Previous tapering attempts | 13 (92.8) | 10 (71.4) | 0.325† |

Data are presented as Mean ± SD, Median [IQR 25%, 75%], or Number (%). Abbreviations:
*OME* Oral Morphine Equivalents, *LTOT* Long-term opioid therapy. \* Independent Sample t-
Test, † Fisher's exact test (Freeman-Halton extension was used for 2 x 3 contingency
tables), ‡ Mann-Whitney U Test, ¥ Missing data for some participants

### Feasibility and acceptability outcomes

Text message delivery was 99.2% successful (778/784). Eight (out of 14, 57.1%) participants in the intervention group confirmed that they had watched the educational video and completed the post-video assessment of tapering self-efficacy. Six other participants did not watch the video, as indicated by the missing post-video assessment. Participants rated the messages as useful (64.2%), easy to understand (78.5%), supportive (71.4%), and encouraging (85.7%) and 78.5% would recommend the intervention to others (Table 3). A posthoc power analysis showed that, with 14 participants in the intervention group, the lower limit of acceptability was 56% (CI = 21.5%) which was above our expected lower limit of 50%. The data completion rate was 85.7% (132 out of 154 assessments).

**Table 3.** Table 3. Acceptability outcomes*.

|  | Median [IQR] | n (%) above neutral† |
| --- | --- | --- |
| Useful | 2.5 [0.75, 3] | 9 (64.2) |
| Helpful | 2 [0.75, 3] | 9 (64.2) |
| Easy to understand | 3 [3, 3] | 11 (78.5) |
| Supportive | 3 [1.75, 3] | 10 (71.4) |
| Bothersome | 1 [-2.25, 3] | 5 (35.7) |
| Encouraging | 3 [2, 3] | 12 (85.7) |
| Would recommend | 3 [1.75, 3] | 11 (78.5) |

\* Assessed using 7-point Likert scale with responses ranging from -3 to 3, with 0 was anchored as neutral. † 12 participants completed the survey, intention-to-treat analysis was performed. *IQR* 25<sup>th</sup> and 75<sup>th</sup> percentiles.

### Potential efficacy outcomes

#### Opioid-tapering self-efficacy

The main effect test showed a higher OTSEQ score in overall across weeks 1 to 4 in the intervention group than the control (estimate [CI80%] = 16.1 [10.9, -], *d* = 0.89). Pairwise contrasts showed a higher OTSEQ score in the intervention group than the control at week 2 (estimate [CI80%] = 9.3 [3.2, -], *d* = 0.49) and week 4 (estimate [CI80%] = 15.6 [8.9, -], *d* = 0.74, Figure 2, Table 4). Moreover, OTSEQ scores were higher after watching the video compared to the baseline in the intervention group (estimate [CI80%] = 9.4 [1.5, -], *d* = 0.27, Figure S2).

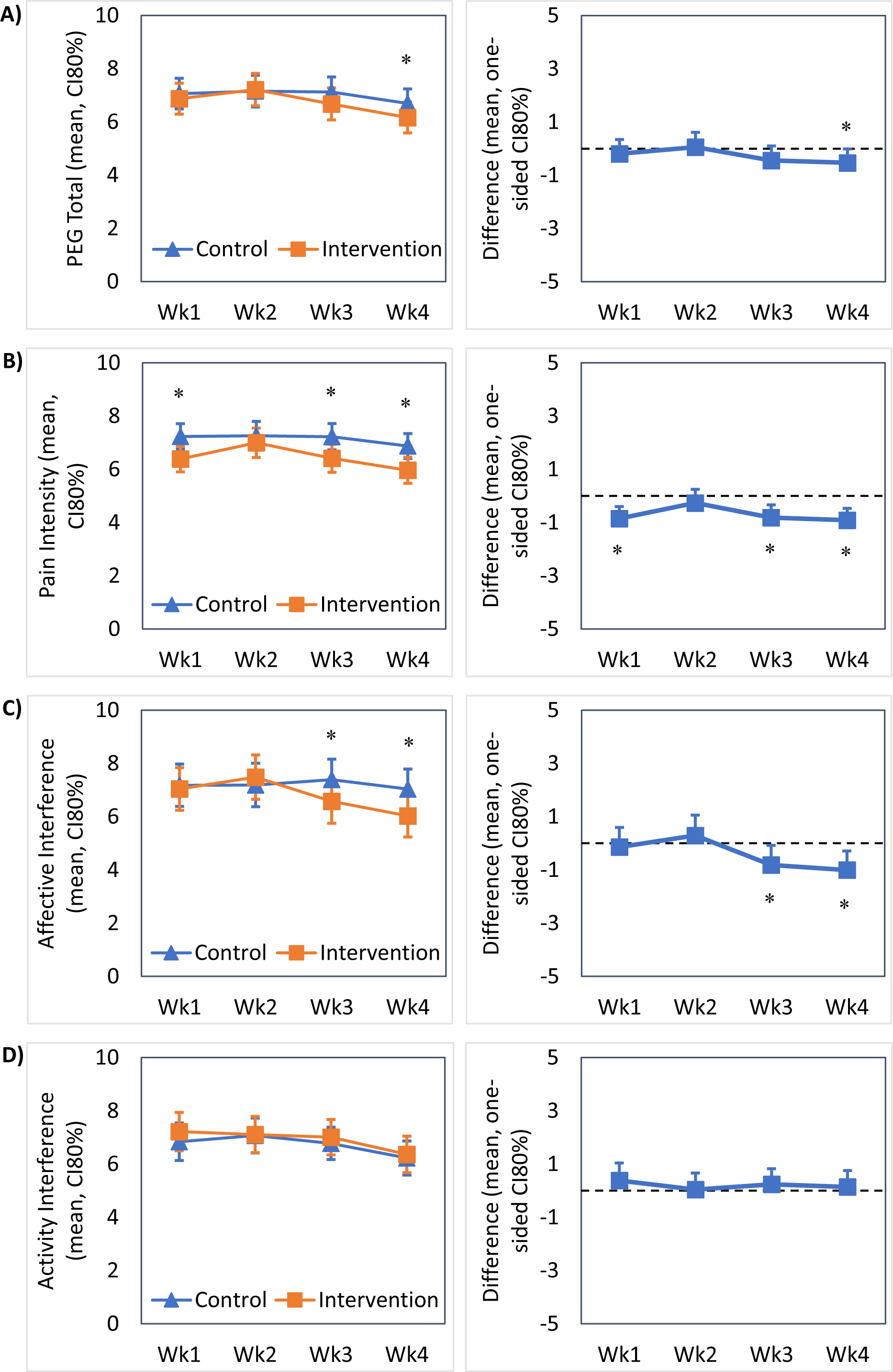

**Table 4.** Comparison of primary and secondary outcomes between the two groups.

|  | Control<br>n = 14 |  | Intervention<br>n = 14 |  |
| --- | --- | --- | --- | --- |
|  | Baseline | Week 4 | Baseline | Week 4 |
| OTSEQ | 62.1 ± 9.7 | 51.0 ± 5.4 | 57.1 ± 8.6 | 66.7 ± 5.7* |
| *PEG total score* | 7.1 ± 0.3 | 6.6 ± 0.4 | 7.1 ± 0.5 | 6.1 ± 0.4* |
| Pain intensity | 6.4 ± 0.3 | 6.8 ± 0.3 | 6.9 ± 0.4 | 5.9 ± 0.3* |
| Pain interference with enjoyment<br>of life | 7.7 ± 0.3 | 7.0 ± 0.5 | 7.2 ± 0.6 | 6.0 ± 0.6* |
| Pain interference with general<br>activity | 7.2 ± 0.4 | 6.2 ± 0.4 | 7.2 ± 0.6 | 6.3 ± 0.5 |
| GAD-2 | 2.7 ± 0.4 | 2.2 ± 0.5 | 2.9 ± 0.5 | 2.3 ± 0.5 |
| PHQ-2 | 2.9 ± 0.4 | 2.4 ± 0.4 | 2.8 ± 0.5 | 2.5 ± 0.4 |
| PSEQ | 29.2 ± 3.3 | 25.9 ± 2.2 | 23.2 ± 4.2 | 29.6 ± 2.4* |
| CAP-6 | 11.9 ± 1.1 | 10.1 ± 1.5 | 12.6 ± 1.8 | 9.2 ± 1.6 |

Data are presented as mean ± standard error. Data of week 4 are estimates from the mixed-effect model output, adjusted for baseline values. All data are presented as Mean ± Standard Error (SE). Abbreviations: *OTSEQ* Opioid Tapering Self-Efficacy Questionnaire, *GAD* Generalized Anxiety Disorder 2-item, *PHQ-2* Patient Health Questionnaire-2, *PSEQ* Pain Self-Efficacy Questionnaire, *CAP-6* Concerns About Pain 6-item. \*Significant difference versus control group at week 4 based on the 80% CI method.

#### PEG scale total score and subscales

The main effect tests showed a lower pain intensity score in overall across weeks 1 to 4 in the intervention group than the control (estimate [CI80%] = -0.8 [-, -0.5], *d* = 0.77), but no difference in PEG scale total score or interference scores. Pairwise contrasts showed that, compared to the control, the intervention group had lower PEG scale total score at week 4 (estimate [CI80%] = -0.5 [-, -0.009], *d* = 0.32), lower pain intensity scores at week 1 (estimate [CI80%] = -0.8 [-, -0.3], *d* = 0.60), week 3 (estimate [CI80%] = -0.8 [-, -0.3], *d* = 0.54), and week 4 (estimate [CI80%] = -0.9 [-, -0.4], *d* = 0.65), and lower affective interference scores at week 3 (estimate [CI80%] = -0.8 [-, -0.08], *d* = 0.35) and week 4 (estimate [CI80%] = -1.0 [-, -0.2], *d* = 0.45, Figure 3, Table 4).

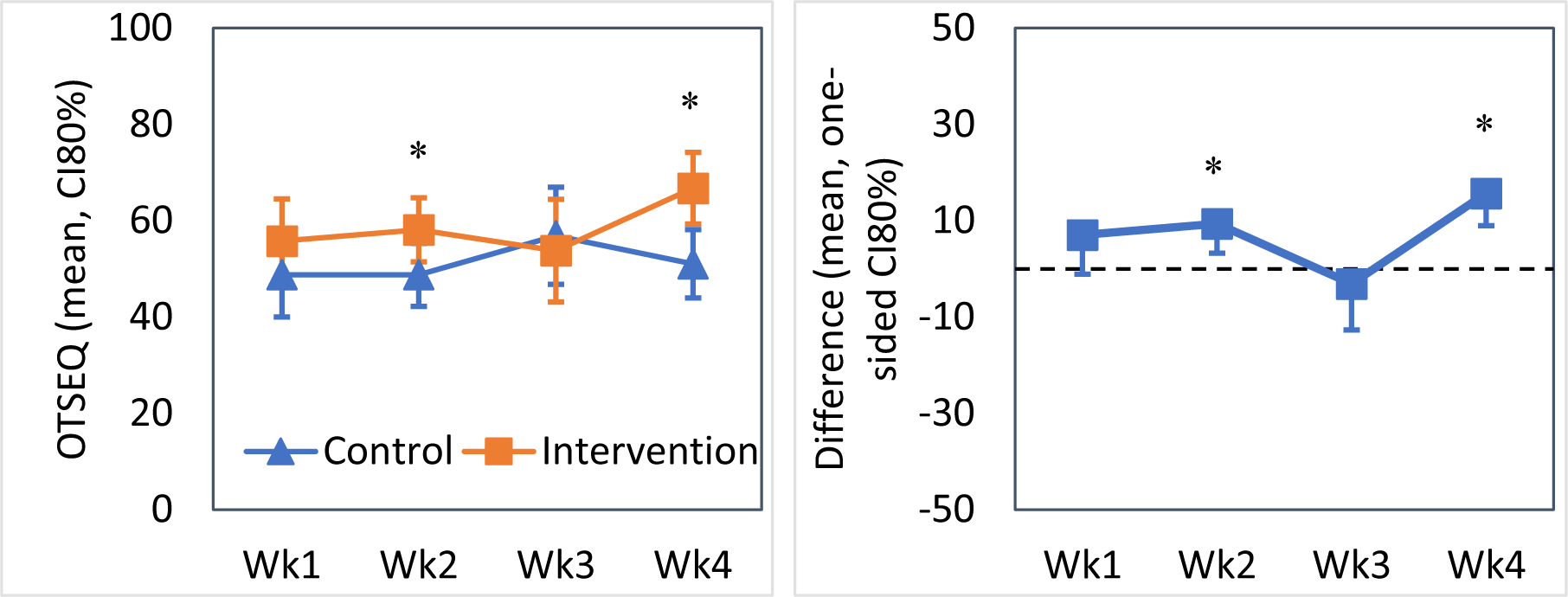

#### Anxiety and Depression

Anxiety and depression scores decreased over time in both the intervention and control groups (see Supplementary Materials for linear models). However, the main effect test showed that anxiety scores were higher in the intervention than the control group overall across weeks 1 to 4 (estimate [CI80%] = 0.7 [0.4, -], *d* = 0.64, Figure 4A). Also, pairwise contrasts showed that, compared to the control, the intervention group had higher anxiety scores at week 2 (estimate [CI80%] = 0.5 [0.2, -], *d* = 0.56) and higher depression scores at week 2 (estimate [CI80%] = 0.7 [0.2, -], *d* = 0.50) and week 3 (estimate [CI80%] = 0.6 [0.004, -], *d* = 0.32). These results were in contrast to the expected direction of the effect.

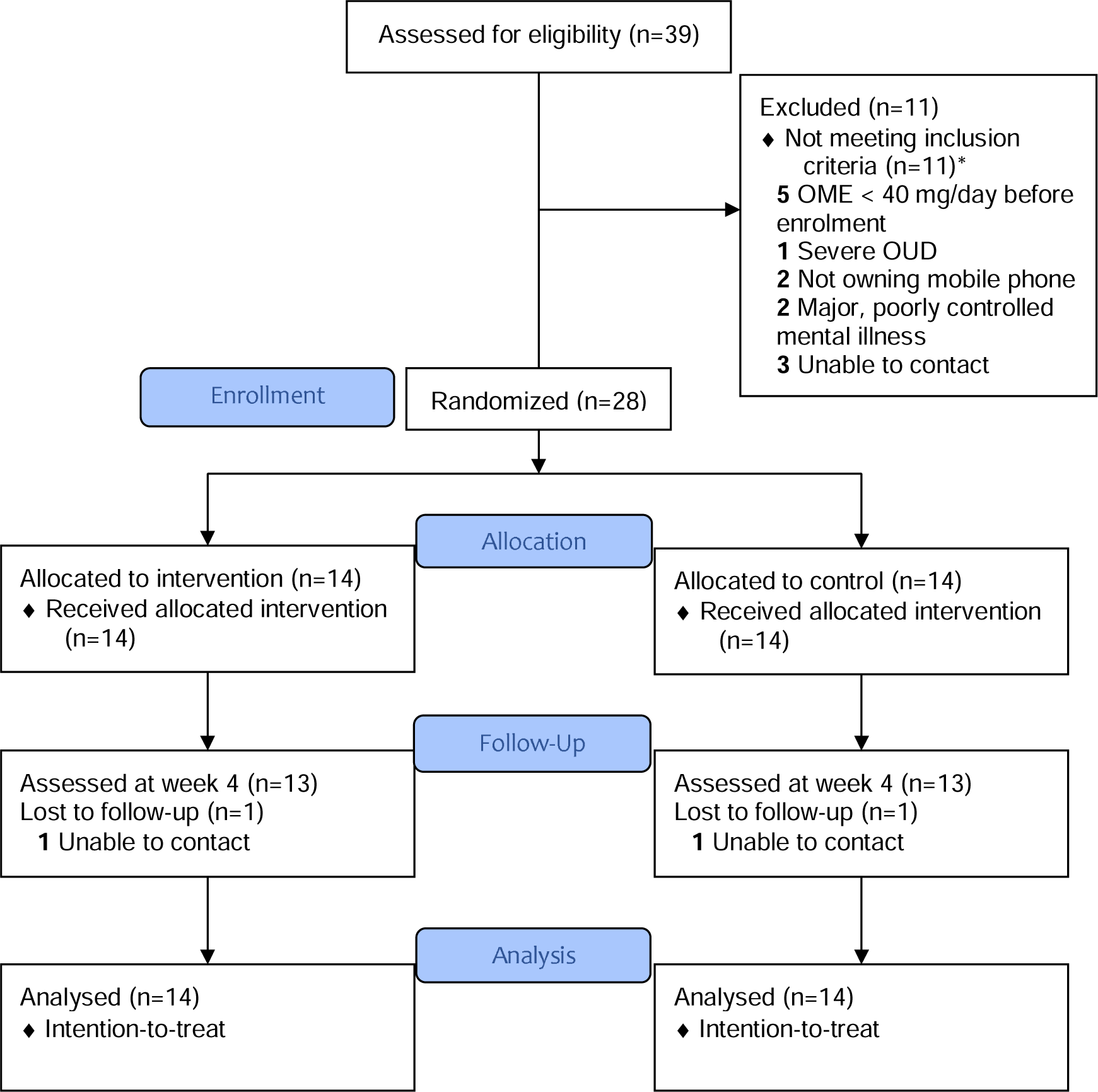

#### Other outcomes

Pain self-efficacy scores at week 4 were higher in the intervention group than the control (estimate [CI80%] = 3.6 [0.7, -], *d* = 0.41, Table 4). At week 4 of the study and compared to baseline, 42.8% (12/28) of the participants had reduced their opioid dose while 14.2% (4/28) had increased and 42.8% (12/28) had no change based on self-reported medications. Opioid dose reduction from baseline to week 4 was not different between the two groups (*p* = 0.892, Table 5). There was no difference between the control and intervention groups in the cumulative number of weeks in which they experienced withdrawal symptoms (median [IQR] = 2 [1, 3] vs. 3 [1.5, 3.5], *p* = 0.530). Satisfaction with care was also not different between the two groups at week 4 (median [IQR] = 6 [4, 7] vs. 6 [6, 7], *p* = 0.878).

**Table 5.** Opioid dose changes from baseline to week 4 between the study groups.

|  | Control<br>n = 14 | Intervention<br>n = 14 | P value |
| --- | --- | --- | --- |
| *Opioid dose changes* |  |  | >0.999* |
| Reduced | 6 (42.8) | 6 (42.8) |  |
| Increased | 2 (14.2) | 2 (14.2) |  |
| No change | 6 (42.8) | 6 (42.8) |  |
| Opioid dose change,<br>OME mg/day | 0 [0, 12.3%] | 0 [0, 10.2%] | 0.458† |

Data are presented as number (%) or median [IQR 25%, 75%]. \* Fisher's exact test with Freeman-Halton extension for 2 x 3 contingency table, † Mann-Whitney U test. *OME* Oral Morphine Equivalent

### Factors associated with opioid tapering self-efficacy

Among the measured factors at baseline, OTSEQ score was positively correlated with autonomy (Spearman’s rho [r] = 0.560), readiness to taper (r = 0.667), social support (r = 0.319), and pain self-efficacy (r = 0.397) and negatively correlated with PEG scale total score (r = -0.545), depression (r = -0.395), and pain catastrophising (r = -0.530) (Figure S9).

### Open-text feedback

#### Preferred frequency, timing, and duration of the text messages

Regarding the frequency of text messaging, 87.5% of the participants reported preferring one to two text messages per day and others preferred less (two or three per week). Most participants (75%) found the timing of text message delivery (mid-morning and mid-afternoon) suitable or had no preference. Also, almost all participants reported that it would be helpful to continue receiving text messages after the four-week intervention for as long as they were tapering their opioid doses (only one participant did not agree).

#### Perceived impact on pain management and feelings about opioid tapering

Participants reported the text messages helped to reinforce and remind them of pain education concepts and pain self-management strategies (“It reminded me that pain is temporary or the feelings are temporary, to exercise, to meditate”). Participants also reported they felt the messages were validating and normalising (“It was brilliant to help me understand what was happening, or what had been happening, to me”) and informative and educational (“Informative ones were interesting, particularly if it contained information you didn’t actually know”). Messages were also found to be supportive and reassuring (“At times tapering opioids I would feel it’s just me, and [that] I was not alone was a helpful message”). Several participants reported the messages helped to keep them motivated (“It made me feel like I could actually succeed, and failure wasn’t an option”) and provided encouragement (“It gave me a sense of achievement”).

#### Barriers to engagement

Many participants said there were no barriers to engagement with the intervention (“Nothing got in the way of engaging with them at all. I always made time, I read the message”). Others said their attitude towards pain management and opioid tapering was not always positive, which made it difficult to engage with the text messages at times (“I was not in the right head space to make this change happen.., but I do believe if I was in the right head space I would have benefited greatly from the messages”). One participant mentioned that the text messages sometimes had the effect of reminding them of the unpleasant aspects of tapering when they had found distraction was more effective. One participant felt the automated nature of the messages was impersonal. However, for another participant, the automated messages felt like genuine social support (“It’s helpful to know that someone has taken the time to send the messages. I thought you were thinking of me”).

## DISCUSSION

The objectives of this pilot RCT were to evaluate the acceptability and feasibility (primary objective) as well as potential efficacy (secondary objective) of an mHealth intervention composed of a psycho-educational video and SMS text message which was co-designed to support people living with chronic pain during opioid tapering. Overall, the results suggest that it is feasible, acceptable, and potentially efficacious to support people living with chronic pain during opioid tapering with this mHealth intervention.

Similar to other studies,^42^ the delivery rate of SMS text messaging was very high. In contrast, many participants in the intervention group did not confirm watching the psycho-educational video. This might be due to low engagement with the video in general and/or due to its delivery method as embedding the video link in the REDCap could reduce its accessibility. Sending the video link to participants directly within the email content (plus using a video thumbnail or an animated GIF) or via text message may increase engagement which can be tested in future studies.

Overall, the study findings suggest that the intervention had a positive impact on opioid-tapering self-efficacy, pain intensity, affective interference, and pain self-efficacy but not on activity interference and pain catastrophising in the short term. Notably, the effect sizes and the estimates of differences became larger over time, indicating that a longer intervention period may be associated with larger and more clinically meaningful effects. The study duration was too short to observe the potential effect of the intervention on opioid dose reduction considering many participants would remain on the same dose for four weeks when being tapered slowly.^36^ It is possible that with longer exposure to the intervention, increasing opioid-tapering self-efficacy may facilitate opioid dose reductions.

Although anxiety and depression symptom severity reduced over the study period in both study groups, scores were higher at some points during the study in the intervention group than the control. The results suggest that the intervention may not have been effective in improving mood in the short term and might have temporarily increased anxiety. It is important to properly investigate the potential negative effects associated with psychological and behavioural interventions.^41^ Therefore, in the definitive trial, we will use The Negative Effects Questionnaire and interview participants to investigate the incidence of a wide range of unwanted events and whether they are attributed to the intervention received. This will provide further insights to revise the intervention characteristics, or use tailoring methods, to reduce any negative effects and maximise benefits.

The qualitative results suggest that the mHealth intervention was generally well-received by participants, with most finding the messages helpful in managing their pain and providing motivation and support. The preferred frequency and timing of the messages varied among participants, with most preferring to receive two messages per day in the morning and evening. Overall, about 78% of the participants mentioned that they would recommend the intervention to others, which is a positive indicator of acceptability.^45^ However, some participants reported that the intervention was bothersome to some extent, and some found the messages to be overwhelming or suggested the need for more personalised support indicating that, similar to other pain interventions, there is likely to be individual variation in the acceptability of mHealth support for opioid tapering necessitating a tailored approach. The findings also suggest that the impact of the messages on pain management and tapering prescription opioids may vary from person to person. While some participants found the messages informative and supportive, others did not feel that the messages had a major influence on how they felt about tapering prescription opioids. Additionally, some participants reported barriers to engaging with the messages, such as negative attitudes. Participant feedback also indicated that engagement with text messages may depend on their readiness to taper. Overall, the qualitative results provide important insights into how patients with CNCP may experience and respond to text message interventions for pain management and opioid tapering. The findings suggest the need for personalised approaches to message delivery, as well as continued support beyond the four-week intervention period. Future research may also explore the potential benefits of more personalised or interactive messaging approaches for pain management and opioid tapering.

This study had strengths and limitations. The study was undertaken at multiple sites, and well randomized. However, the sample size was small and recruitment had to cease before reaching the planned sample size. The slow recruitment rate might be due to lack of interest, strict inclusion and exclusion criteria, or other reasons and warrants further investigation. Although we achieved the sample size that was required for most of the study aims, these limitations can potentially affect the validity and generalizability of the study findings.

## CONCLUSION

Prescription opioid tapering can be fraught with challenges and distress for people living with chronic pain. Often, behavioral support is lacking, and scalable, low-cost, and low-burden options are needed to provide daily support to people living with chronic pain throughout their tapering process. This pilot study shows that it is feasible, acceptable, and potentially efficacious to support patients with CNCP during opioid tapering with a co-designed psycho-educational video and SMS text messaging intervention. This intervention can potentially increase self-efficacy in tapering opioids and improve pain intensity and interference. Considering the promising results of this pilot RCT and based on feasibility findings, a definitive trial has been designed and initiated. The duration of the intervention has been increased to 12 weeks based on feedback received from participants in this pilot RCT. Also, the frequency, interval, and number of questionnaires in the assessments have been reduced to minimise the burden on participants and increase the data completion rate. The definitive trial will recruit participants directly from the community with no direct referral being required. This may help to conduct more inclusive pain research,^25^ and will enable us to investigate the barriers and facilitators of implementing this digital support on a large scale and to prepare strategies accordingly.

## Authors Contributions

PG and CEAJ conceptualized the interventions and acquired funding and should be considered joint senior authors. AG, MRM, AGM, LD, AS, MF, BDD, PG, and CEAJ contributed to the design of the study protocol. MRM coordinated the recruitment and intervention delivery. CEAJ, PG, TB, AA, MC, and IH supervised the recruitment at the study sites. AG, MRM, and AGM collected the data. AG performed the statistical analysis and drafted the manuscript. All authors contributed to the interpretation of the results, critically reviewed and edited the manuscript, and approved the final submitted version.

## Funding

This research was supported by a philanthropic gift to The University of Sydney from the Ernest Heine Family Foundation. The study funder and sponsor had no role in the study design, data collection, analysis, and interpretation, or in preparing the final report. BDD acknowledges support from the National Institutes of Health NIDA K24-DA053564.

## Conflicts of Interest

Dr. Darnall is Chief Science Advisor at AppliedVR, and her consulting role with this company (personal fees) is unrelated to the current research. Dr. Darnall receives royalties for four pain treatment books she has authored or co-authored. She is the current principal investigator for two pain research awards from the Patient-Centered Research Outcomes Research Institute and two NIH pain research grants. She serves on the Board of Directors for the American Academy of Pain Medicine, the Board of Directors for the Institute for Brain Potential, and the Medical Advisory Board for the Facial Pain Association. Dr. Darnall is a scientific member of the NIH Interagency Pain Research Coordinating Committee, a former scientific member of the Centers for Disease Control and Prevention Opioid Workgroup (2020–2021), and a current member of the Pain Advisory Group of the American Psychological Association. Other Authors have no conflict of interest.

## Data Availability

Individual data produced in the present study are not available to maintain patient confidentiality. SAS codes and output tables and additional analysis may be provided upon reasonable request to the authors.

## Notes

### Clinical Trial

ACTRN12621000795897

### Clinical Protocols

https://bmjopen.bmj.com/content/12/4/e057174

https://anzctr.org.au/Trial/Registration/TrialReview.aspx?id=381757&isReview=true

### Author Declarations

The study was approved by the Northern Sydney Local Health District Research and Ethics Committee (ID number 2020/ETH03288) and pre-registered with the Australian New Zealand Clinical Trials Registry (registration number ACTRN12621000795897).

