## Supplementary Materials for "A digital health intervention to support patients with chronic pain during prescription opioid tapering: a pilot randomised controlled trial"

**Supplementary File**

**METHODS**

Details of the study methods are described in the published study protocol.^4^ The study protocol was submitted for publication before the enrolment of the first participant into the study. Changes to the study protocol before trial commencement (recruitment) are listed with the details in the Australian New Zealand Clinical Trials Registry (registration number ACTRN12621000795897). These include changing 60 mg OME to 40 mg OME in the inclusion criteria and changes in the study questionnaires. Other than adding new sites to the study, there was no important change to the methods after trial commencement (recruitment).

Table S1: Data collection plan

|  | Description | Baseline | Week 1 | Week 2 | Week 3 | Week 4 |
| --- | --- | --- | --- | --- | --- | --- |
| Demographics – Clinical data | Age, Gender, Socioeconomic, Pain, Comorbidities, Medications, Tapering history | X |  |  |  |  |
| Autonomy | Single-item scale | X |  |  |  |  |
| Physician-patient relationship | PDRQ-9 | X |  |  |  |  |
| Social support | OSSS-3 | X |  |  |  |  |
| Readiness to taper | Single-item scale | X |  |  |  |  |
| Expectancy | Pain, Enjoyment, Activity, Mood | X |  |  |  |  |
| Self-efficacy in tapering | Single-item OTSEQ^a^ | X | X | X | X | X |
| Pain | 3-item PEG scale | X | X | X | X | X |
| Emotional functioning | PHQ-2 and GAD-2 | X | X | X | X | X |
| Opioid dose, OME | Self-report | X | X | X | X | X |
| Withdrawal symptoms | Self-report |  | X | X | X | X |
| Pain cognitions | CAP-6 and PSEQ | X |  |  |  | X |
| Current care |  | X |  |  |  | X |
| Satisfaction with care | Single-item scale |  |  |  |  | X |
| Acceptability | Feedback survey, interview |  |  |  |  | X |
| Feasibility | Recruitment, dropout, delivery |  |  |  |  | X |

Abbreviations: PEG: Pain Enjoyment of life General activity scale; PHQ-2: Patient Health Questionnaire-2; GAD-2: Generalized Anxiety Disorder-2; OME: Oral Morphine Equivalents; CAP-6: Concerns about Pain Scale; PSEQ: Pain Self-Efficacy Questionnaire; OSSS-3: Oslo Social Support Scale. ^a^ OTSEQ was repeated after watching the video.

The following feedback survey was used at week 4 to capture the likelihood of recommending the intervention to others, perceived usefulness of the intervention, levels of engagement with the intervention, barriers and facilitators of engagement, messages readability, and feasibility of frequency and timing of the messages.^3,5^

**Feedback Survey**

We would like to thank you for your participation in this study. We would like to invite you to take part in our feedback survey so that we can evaluate the text message intervention. This survey should take no more than 10 minutes of your time. Thank you for your continued participation.

**Instructions:** We would be grateful for feedback on your experiences of receiving text messages to provide additional support during opioid tapering. Please feel free to share your thoughts openly and honestly. We encourage you to share your thoughts on whether or not the text messages helped you and, if so, how. Please also share anything else about your experience that you think might be relevant.

1. Please write your feedback here:

2. Please use the following scales to describe your experience of receiving text messages to support opioid tapering over the past 4 weeks. You can move the slider across the scale.

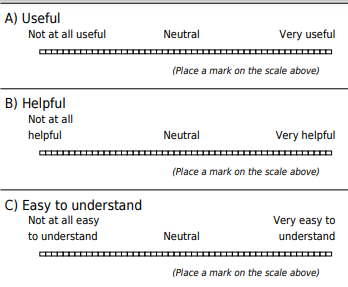

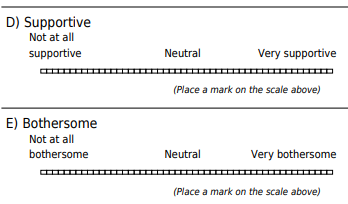

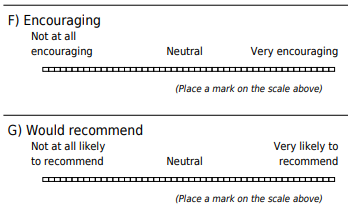

3. Please answer the following open-ended questions as honestly as you can:

A) You received 2 text messages per day as part of this study. How many text messages per day or per week do you think would be ideal? (Please specify number per day or per week).

B) You received text messages two times each day. What time/s of the day are NOT ideal for receiving supportive text messages?

C) Did receiving the text messages influence how you manage your pain? If so, how?

D) Did receiving the text messages influence how you felt about tapering prescription opioids? If so, how?

E) Which factors did you see as barriers to your engagement with the text messages?

F) You received 2 text messages per day for 4 weeks as part of this study. Do you think it would be helpful to keep receiving text messages beyond this period? Yes No

F 1) How long (weeks or months) would you suggest the text messaging continue?

F 2) How many SMS per day or per week would you suggest the text messaging continue? (please specify per day or per week in your answer)

4. Please use the scales provided to indicate your engagement with text messages:

A) How often did you take up advice you received in the text messages?

Never / Almost never / Sometimes / Most of the times / Always

B) Did you find that the advice given helped you to manage your pain better?

Not at all / Not really / Neutral / Somewhat / Absolutely

C) Did you think that the messages helped you to reduce your prescription opioid dose?

Not at all / Not really / Neutral / Somewhat / Absolutely

D) Were you satisfied with the text message intervention received?

Not satisfied at all / Not very satisfied / Neutral / Somewhat satisfied / Extremely satisfied

5. Do you have anything else to add about your experience with receiving this SMS support? Please write your feedback here:

*Opioid-Tapering Self-Efficacy Questionnaire*

A one-item scale of general self-efficacy to taper prescription opioid (OTSEQ, Opioid-Tapering Self-Efficacy Questionnaire) was developed for the study using Bandura’s self-efficacy theory and guides for constructing self-efficacy scales.^1^ The scale was evaluated for face validity by a panel of clinicians, and cognitive debriefing was conducted by interviewing six patients with CNCP who had experienced opioid tapering.

**Instructions:** Your doctor has recommended you to gradually reduce the dose of the prescription opioid medication you are currently using for pain. This scale measures your confidence in your ability to reduce prescription opioid medication. Please indicate how confident you are at the present time by selecting one number on the scale from 0 (indicating not at all confident) to 100 (indicating completely confident).

1. How confident are you at the present time that you can reduce your dose of opioid medication? (0 = Not at all confident, 100 = Completely confident)

⃝ 0	⃝ 10	⃝ 20	⃝ 30	⃝ 40	⃝ 50	⃝ 60	⃝ 70	⃝ 80	⃝ 90	⃝ 100

*Exploring factors associated with opioid tapering self-efficacy*

To evaluate expectancy participants completed the following scale.

What would you expect to happen to the following as a result of reducing your dose of opioid medications over the next 4 weeks?

Responses: ⃝ Better ⃝ No change ⃝ Worse ⃝ Not sure

1. Pain

2. Enjoyment of life

3. General activity

4. Mood Better

To evaluate autonomy participants completed the following scale.

To what extent was it your decision to reduce your dose of opioid medications?

⃝ Completely my decision

⃝ Shared decision

⃝ Not at all my decision

**Statistical analysis**

*Sample size*

To assess whether the intervention is acceptable to 70% of the patients with a 20% precision rate, 18 participants were needed for the intervention arm. To evaluate the potential efficacy of the intervention, 12 participants were needed in each group assuming a medium standardised effect size (Cohen’s *d* = 0.5) and using the 80% one-sided confidence interval (CI) approach, which is recommended for pilot trials.^2^ To gather data to obtain estimates and calculate sample size for the future definitive trial, a sample size of 15 has been recommended for each study arm for a medium standardised effect size.^2^ It was expected that there would be a 10% loss to follow-up during the study period. Accordingly, the sample size was set at 20 participants for each study arm to address the primary and secondary objectives of the study.

*Statistical methods*

Descriptive statistics were used for reporting demographic and clinical characteristics as well as for feasibility and acceptability measures. Independent sample t-Test, Man-Whitney U Test and Fisher’s Exact Test were used to compare the two groups regarding baseline and demographic characteristics. The linear mixed-effects model was used to analyse outcomes of potential efficacy. Model statement and parameters were determined based on the fit statistics and residuals distribution. Normal distribution of the residuals was tested. If departing from normality, data was transformed if appropriate to approximate residuals to normality. Missing data were imputed by the mixed-model analysis without any further ad hoc imputation.

First, the main effect of the group was tested to estimate the overall difference in outcomes between the two groups including all time points after randomisation. To account for the correlation between repeated measures, a random intercept was included at the subject level. Then, group by time interaction was included in the model to assess if and how the effect might have changed over time. Considering the small sample size and to control for any baseline differences between groups, we tested if baseline adjustment can improve the precision of the estimates. We expected to observe the maximum potential effect at week 4. In addition, pairwise contrasts were done to explore comparing the outcomes between the two groups at each week of the study.

According to the pre-registered analysis plan,^4^ we used the one-sided 80% CI method in this pilot study. With this approach, we were interested in whether the difference estimates were larger or smaller than zero (depending on the predicted direction of the effect), and did not aim to formally undertake hypothesis testing procedures to prove the efficacy of the intervention.^2^ Hence, no correction was done for multiple comparisons for this pilot study. Cohen’s *d* (effect size) was calculated based on the estimates and standard errors.

Fisher’s exact test and Mann-Whitney U test were used for comparing opioid dose change (reduced, increased, no change) and percentage of reduction between the two groups, respectively. The cumulative incidence of withdrawal symptoms and satisfaction were compared using the Mann-Whitney U test. These analyses were done based on intention-to-treat analysis and missing data were imputed using the last observation carried forward method. Correlation analyses were done to find factors associated with opioid tapering self-efficacy.

**Results**

*Opioid-tapering self-efficacy*

There was a significant main effect of group on OTSEQ score (F = 7.13, P = 0.013). No significant group by time interaction was observed (F = 1.75, P = 0.165). Linear model showed an increase in OTSEQ over time in the intervention group (positive slope; estimate = 3.0, CI80% = 0.6 to 5.4) but no such trend in the control group (slope estimate = 1.4, CI80% = -0.8 to 3.6). OTSEQ scores were higher, in overall, in the intervention group than the control (Estimate [one-sided CI80%] = 9.1 [3.3, -], *d* = 0.48), but the main effect of group in the model was not significant (F = 1.79, P = 0.188) nor was group by time interaction (F = 0.41, P = 0.527). Pairwise contrasts showed a higher OTSEQ score in the intervention compared to the control group at week 3 (Estimate [one-sided CI80%] = 8.2 [2.3, -], *d* = 0.44) and week 4 (Estimate [one-sided CI80%] = 9.8 [3.9, -], *d* = 0.53).

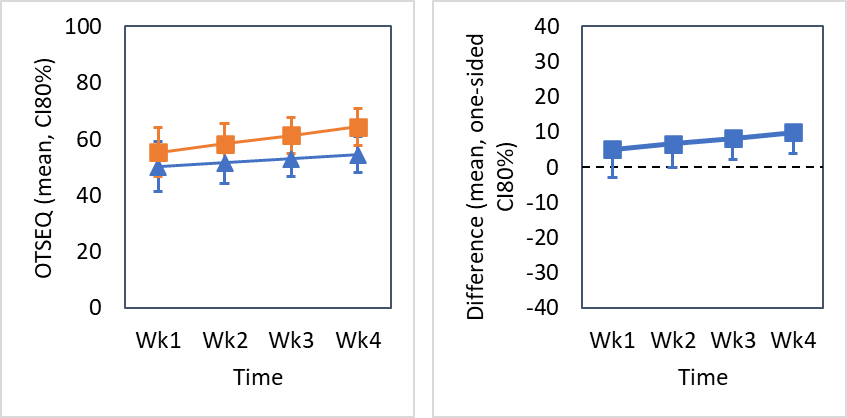

Figure S1: Linear changes in OTSEQ from week 1 to 4

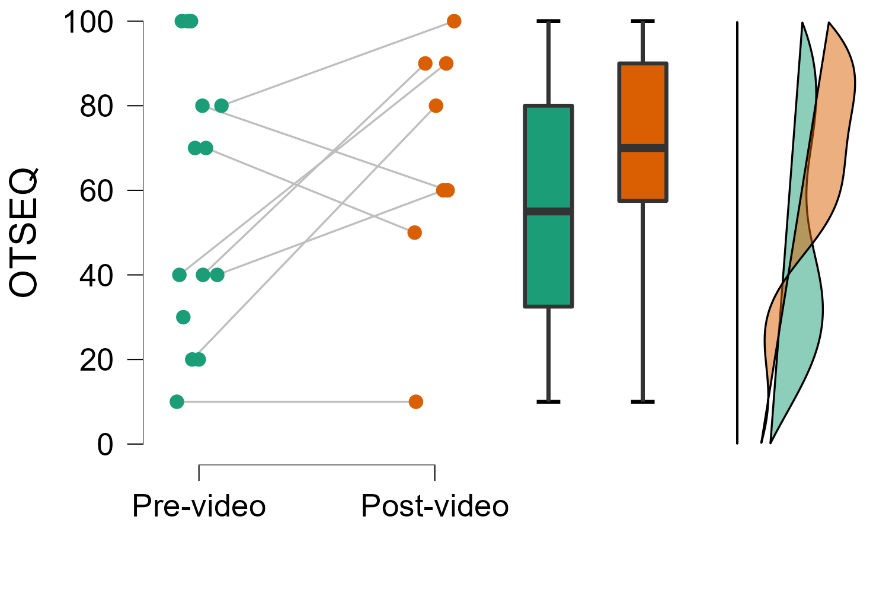

Figure S2. Opioid tapering self-efficacy (OTSEQ) scores before and after watching the psychoeducational video (intervention group only).

*PEG total score*

There was no significant main effect of group on PEG total score (F = 0.37, P = 0.550) and no group by time interaction (F = 0.37, P = 0.771). Linear model showed a decrease in PEG total score over time in the intervention group (negative slope; estimate = -0.2, CI80% = -0.4 to -0.1) but no such trend in the control group (slope estimate = -0.1, CI80% = -0.2 to 0.04). Linear model showed no significant main effect of group on PEG total score (F = 0.11, P = 0.738) and no group by time interaction (F = 0.90, P = 0.349). PEG total score was not lower in the intervention than the control group (Estimate [one-sided CI80%] = -0.1 [-, 0.2]). Pairwise contrasts showed no lower PEG total score in the intervention compared to the control group at week 1 to 4 (one-sided CI80% upper limit at all weeks was larger than zero).

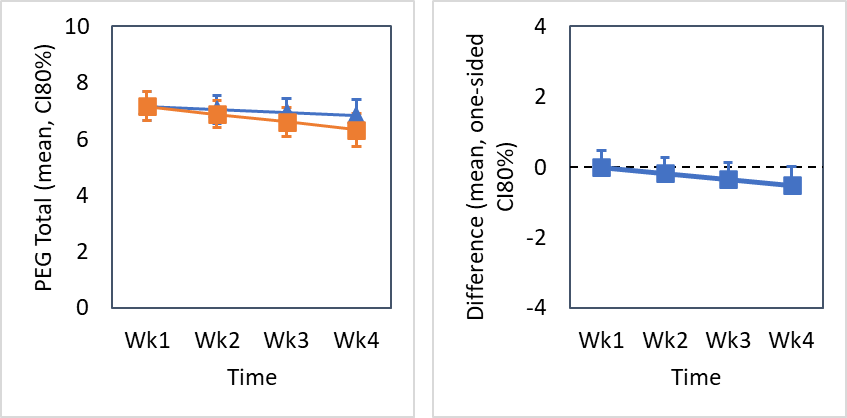

Figure S3: Linear changes in PEG total score from week 1 to 4

*PEG – Pain Intensity subscale*

There was a significant main effect of group on pain intensity (F = 4.42, P = 0.046). No significant group-by-time interaction was observed (F = 0.77, P = 0.517). Linear model showed a decrease in pain intensity over time in the intervention group (negative slope; estimate = -0.1, CI80% = -0.3 to -0.01) but no such trend in the control group (slope estimate = -0.1, CI80% = -0.2 to 0.04). Pain intensity was lower, in overall, in the intervention group than the control (Estimate [one-sided CI80%] = -0.6 [-, -0.2], *d* = 0.54), but the main effect of group in the model was not significant (F = 2.07, P = 0.157) nor was group by time interaction (F = 0.17, P = 0.683). Pairwise contrasts showed a lower pain intensity in the intervention compared to the control group at week 1 (Estimate [one-sided CI80%] = -0.5 [-, -0.1], *d* = 0.42), week 2 (Estimate [one-sided CI80%] = -0.6 [-, -0.2], *d* = 0.53), week 3 (Estimate [one-sided CI80%] = -0.7 [-, -0.3], *d* = 0.56), and week 4 (Estimate [one-sided CI80%] = -0.7 [-, -0.3], *d* = 0.53).

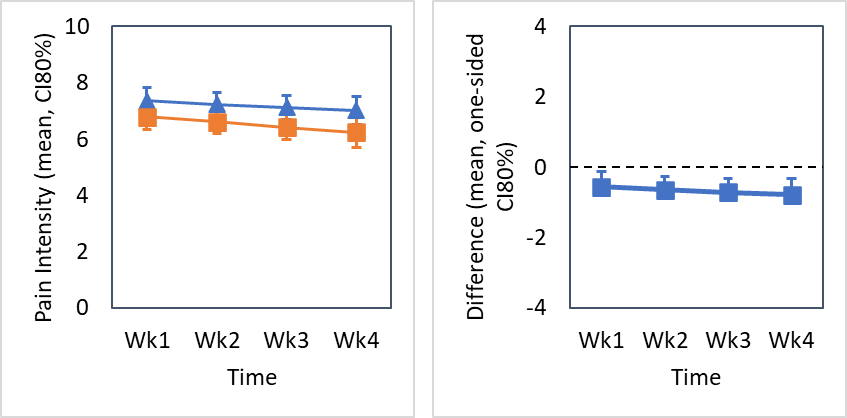

Figure S4: Linear changes in pain intensity from week 1 to 4

*PEG – Pain Interference with Enjoyment of Life subscale*

There was no significant main effect of group on affective interference (F = 0.71, P = 0.406) and no group by time interaction (F = 0.90, P = 0.444). Linear model showed a decrease in affective interference score over time in the intervention group (negative slope; estimate = -0.4, CI80% = -0.6 to -0.1) but no such trend in the control group (slope estimate = -0.02, CI80% = -0.2 to 0.1). Linear model showed no significant main effect of group on affective interference score (F = 0.05, P = 0.821) and no group by time interaction (F = 2.96, P = 0.092). Affective interference score, averaged across all time points, was not lower in the intervention than the control group (Estimate [one-sided CI80%] = -0.1 [-, 0.4]). Pairwise contrasts showed lower affective interference score in the intervention compared to the control group at week 4 (Estimate [one-sided CI80%] = -0.9 [-, -0.2], *d* = 0.42).

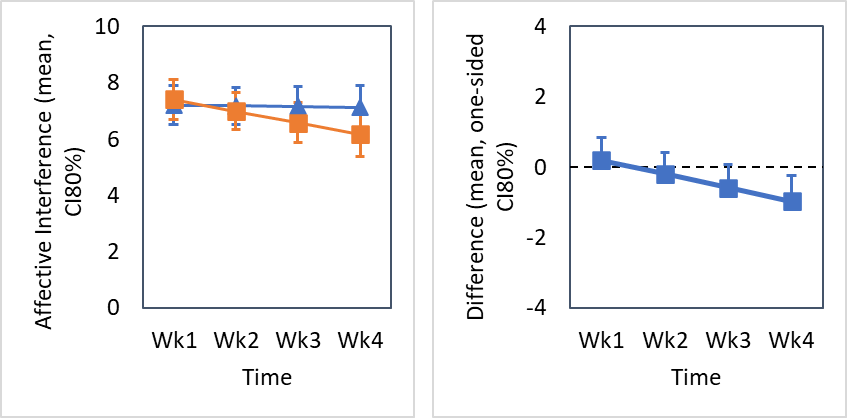

Figure S5: Linear changes in affective interference from week 1 to 4

*PEG – Pain Interference with General Activity subscale*

There was no significant main effect of group on activity interference (F = 0.15, P = 0.698) and no group by time interaction (F = 0.08, P = 0.968). Linear model showed a decrease in activity interference score over time in the intervention group (negative slope; estimate = -0.2, CI80% = -0.4 to -0.02) but no such trend in the control group (slope estimate = -0.2, CI80% = -0.4 to 0.01). Linear model showed no significant main effect of group on activity interference score (F = 0.15, P = 0.704) and no group by time interaction (F = 0.05, P = 0.819). Activity interference score, averaged across all time points, was not lower in the intervention than the control group (Estimate [one-sided CI80%] = -0.1 [-, 0.4]). Pairwise contrasts showed no lower activity interference score in the intervention compared to the control group at week 1 to 4 (one-sided CI80% upper limit at all weeks was larger than zero).

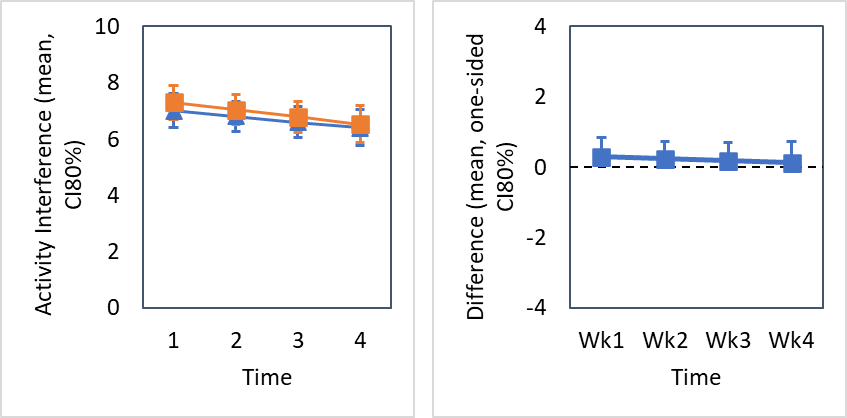

Figure S6: Linear changes in activity interference from week 1 to 4

*Anxiety*

There was a significant main effect of group on anxiety (F = 4.79, P = 0.038), however the effect was in opposite of the expected direction. There was no group by time interaction (F = 0.56, P = 0.640). Pairwise contrasts showed higher anxiety score in the intervention compared to the control group at week 2 (Estimate [one-sided CI80%] = 0.5 [0.2, -], d = 0.56) which was observed also with the two-sided test (CI80% = 0.07, 0.9). Linear model showed a decrease in anxiety over time in both the intervention (negative slope; estimate = -0.2, CI80% = -0.4 to -0.08) and control group (negative slope; estimate = -0.2, CI80% = -0.3 to -0.05). Linear model showed no significant main effect of group on anxiety (F = 0.27, P = 0.608) and no group by time interaction (F = 0.07, P = 0.793). Anxiety score, averaged across all time points, was not lower (or higher) in the intervention than the control group (Estimate [one-sided CI80%] = 0.2 [-0.1, -]). Pairwise contrasts showed no lower (or higher) anxiety score in the intervention compared to the control group at week 1 to 4 (one-sided CI80% upper limit at all weeks was larger than zero and two-sided CI80% included zero).

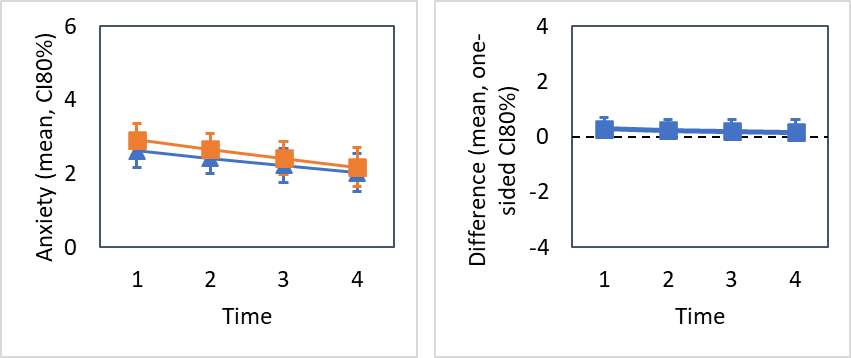

Figure S7: Linear changes in anxiety from week 1 to 4

*Depression*

There was no significant main effect of group on depression (F = 0.35, P = 0.557) and no group by time interaction (F = 0.95, P = 0.422). Pairwise contrasts in the marginal model showed higher depression score in the intervention compared to the control group at week 2 (Estimate [one-sided CI80%] = 0.7 [0.2, -], *d* = 0.50) and week 3 (Estimate [one-sided CI80%] = 0.6 [0.004, -], *d* = 0.32) which was observed also with the two-sided test but only at week 2 (CI80% = 0.02, 1.3). Linear model showed a decrease in depression over time in both the intervention (negative slope; estimate = -0.2, CI80% = -0.3 to -0.04) and control group (negative slope; estimate = -0.1, CI80% = -0.3 to -0.03). Linear model showed no significant main effect of group on depression (F = 0.68, P = 0.413) and no group by time interaction (F = 0.03, P = 0.870). Depression score, averaged across all time points, was not lower (or higher) in the intervention than the control group (Estimate [one-sided CI80%] = 0.4 [-0.01, -], two-sided CI80% = -0.2, 1.0). Pairwise contrasts showed no lower (or higher) depression score in the intervention compared to the control group at week 1 to 4 (one-sided CI80% upper limit at all weeks was larger than zero and two-sided CI80% included zero).

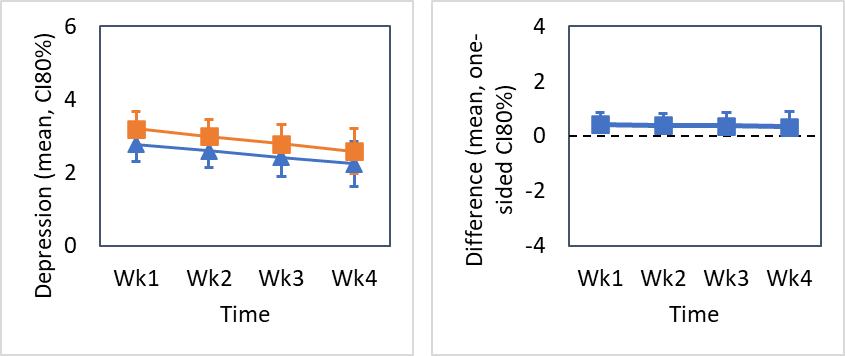

Figure S8: Linear changes in depression from week 1 to 4

*Other outcomes*

There was no significant main group effect on pain-self efficacy (F = 1.16, P = 0.293) or pain catastrophising (F = 0.18, P = 0.674).

**Factors associated with opioid tapering self-efficacy**

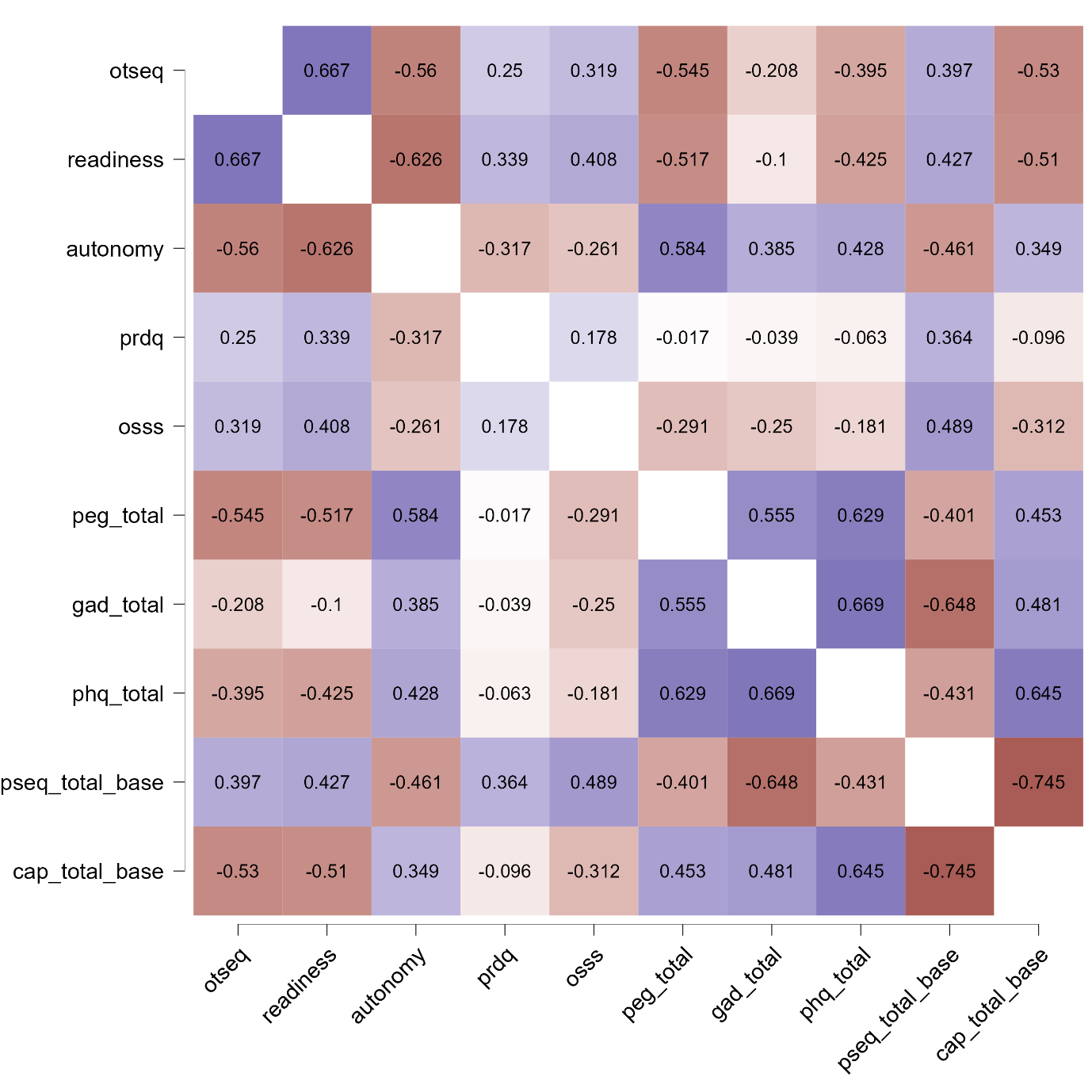

Figure S9. Spearman's rho heatmap of factors associated with opioid tapering self-efficacy at baseline. OTSEQ Opioid Tapering Self-Efficacy Questionnaire, GAD Generalized Anxiety Disorder 2-item, PHQ-2 Patient Health Questionnaire-2, PSEQ Pain Self-Efficacy Questionnaire, CAP-6 Concerns About Pain 6-item, PDRQ Patient-Doctor Relationship Questionnaire, OSSS Oslo Social Support Scale.

*Patients’ baseline expectations regarding outcomes following opioid tapering and its association with opioid tapering self-efficacy.*

Table S2: Patients’ baseline expectations* regarding outcomes following opioid tapering.

|  | Control  n = 14 | Intervention  n = 14 | P value† |
| --- | --- | --- | --- |
| Pain |  |  | 0.304 |
| Better | 3 (21.4) | 0 (0) |  |
| No change | 1 (7.1) | 3 (21.4) |  |
| Worse | 6 (42.8) | 7 (50) |  |
| Not sure | 4 (28.5) | 4 (28.5) |  |
| Enjoyment of life |  |  | 0.956 |
| Better | 3 (21.4) | 3 (21.4) |  |
| No change | 2 (14.2) | 3 (21.4) |  |
| Worse | 5 (35.7) | 5 (35.7) |  |
| Not sure | 4 (28.5) | 3 (21.4) |  |
| General activity |  |  | 0.790 |
| Better | 2 (14.2) | 1 (7.1) |  |
| No change | 2 (14.2) | 4 (28.5) |  |
| Worse | 5 (35.7) | 4 (28.5) |  |
| Not sure | 5 (35.7) | 5 (35.7) |  |
| Mood |  |  | 0.348 |
| Better | 3 (21.4) | 1 (7.1) |  |
| No change | 1 (7.1) | 4 (28.5) |  |
| Worse | 5 (35.7) | 6 (42.8) |  |
| Not sure | 5 (35.7) | 3 (21.4) |  |

* Expectation was assessed before randomisation

† Fisher’s exact test (Freeman-Halton extension was used for 2 x 4 contingency tables)

Overall, patients who expected worse outcomes following opioid tapering had lower tapering self-efficacy at baseline.

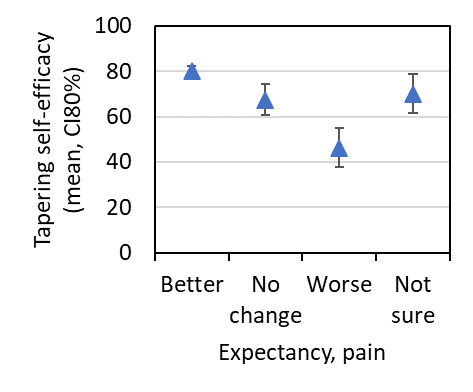

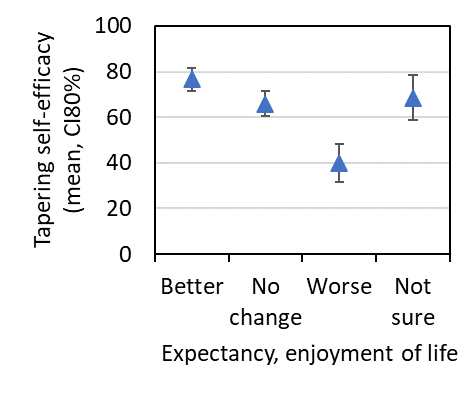

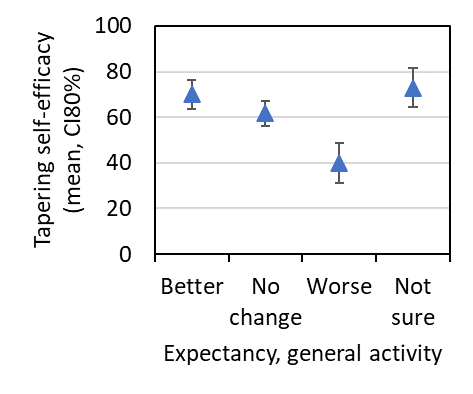

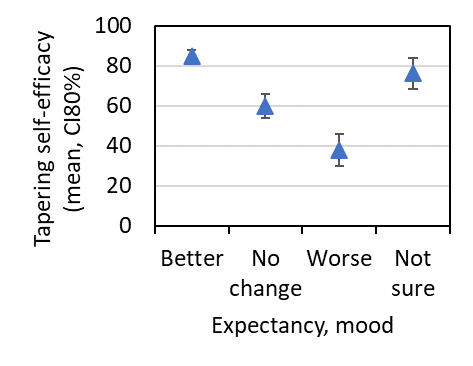

Figure S10. Association between patients’ baseline expectations regarding outcomes following opioid tapering and baseline tapering self-efficacy.

Table S3: Healthcare visits for pain management during the study period (based on self-report)

|  | Control  n = 13 | Intervention  n = 11 | P value† |
| --- | --- | --- | --- |
| Specialist pain physician | 9 (69.2) | 8 (72.7) | > 0.999 |
| Psychologist | 7 (53.8) | 5 (45.4) | > 0.999 |
| Psychiatrist | 2 (15.3) | 2 (18.1) | > 0.999 |
| Physiotherapist | 6 (46.1) | 2 (18.1) | 0.210 |
| Occupational therapist | 2 (15.3) | 0 (0) | 0.481 |
| Nurse | 2 (15.3) | 2 (18.1) | > 0.999 |

† Fisher’s exact test
